# Gray matter networks associated with cognitive deficit in ADHD across adolescence and adulthood

**DOI:** 10.1101/2020.04.22.20059808

**Authors:** Jingyu Liu, Kuaikuai Duan, Wenhao Jiang, Kelly Rootes-Murdy, Gido Schoenmacker, Jan K. Buitelaar, Martine Hoogman, Jaap Oosterlaan, Pieter J. Hoekstra, Dirk J. Heslenfeld, Catharina A. Hartman, Vince D. Calhoun, Alejandro Arias-Vasquez, Jessica A. Turner

**Author notes:** Co-first author (Jingyu Liu, Kuaikuai Duan). Corresponding author: Dr. Jingyu Liu.

## Abstract

Attention-deficit/hyperactivity disorder (ADHD) is a childhood-onset neuropsychiatric disorder, and its existence in adulthood is well established. Beyond symptoms of inattention and hyperactivity/impulsivity, patients commonly present with impairments in cognition. How neuronal underpinnings of symptoms and cognitive deficits differ across adolescence and adulthood is not clear. In this cross sectional study, we investigated gray matter of two cohorts, 486 adults and 508 adolescents, each including participants with ADHD and healthy controls. Independent component analysis was applied to the gray matter of each cohort, separately, to extract cohort specific networks. Then, we identified gray matter networks associated with symptoms, working memory and/or diagnosis in each cohort, and projected them onto the other cohort for comparison. Two components in the inferior, middle/superior frontal regions identified in adults and one component in the insula and inferior frontal region identified in adolescents were significantly associated with working memory deficits in both cohorts. One component in bilateral cerebellar tonsil and culmen identified in adults and one component in left cerebellar region identified in adolescents were significantly associated with inattentive symptoms in both cohorts. All these components presented significant or nominal level of gray matter reduction for ADHD patients in adolescents, but only one showed nominal reduction for patients in adults. Our findings suggest gray matter reduction may not be a sensitive marker for persist ADHD. However, the patterns of certain brain regions are associated with deficits in working memory or attention persistently from childhood into adulthood, which might help understand the mechanism of disease persistence.

## Introduction

Attention-deficit/hyperactivity disorder (ADHD) is a childhood-onset neuropsychiatric disorder characterized by attentional problems, and/or hyperactivity and impulsivity. Beyond the symptoms, patients with ADHD also present with heterogeneous impairments in multiple domains of cognition ^1-6^. Recently ADHD persistence into adulthood has been observed in 15% to 50% of cases ^7, 8^, depending on whether or not counting partial remission ^9, 10^. Decades of studies of brain structure and function have accumulated adequate evidence indicating the implication of several key brain regions, highlighting fronto-striatal, fronto-parietal and fronto-cerebellar networks ^11^. The structure and function of these networks change with age ^12, 13^. Thus, their dynamics might be related to the persistence or remission of the disorder ^11, 14^. As reviewed by Sudre et al., multiple processes including ‘neural normalization’, ‘neural reorganization’, and ‘fixed anomaly’ might occur in different brain networks during the progress of ADHD ^14^. A pressing need is to investigate neuronal underpinnings of ADHD with emphases on the relationship of neuronal alterations of patients in childhood, adolescence and adulthood.

In this study we focused on gray matter (GM) in relation to ADHD symptoms and cognitive deficits. Studies of children and/or adolescents have reported GM reduction in widespread brain regions, while the most common effects are in the subcortical regions ^13, 15, 16^ and cerebellum ^17^, followed by regions in the frontal, parietal, temporal cortex ^18-20^. How the GM reduction links to symptoms or cognitive deficits is not entirely clear. Castellanos et al. showed that in children and adolescent patients with ADHD, GM volumes of frontal and temporal lobes, caudate, and cerebellum were negatively correlated with the overall score of illness severity, and particularly, attention problems ^20^. However, Jacobson et al. showed that GM reduction in frontal, temporal and parietal lobes were more associated with hyperactive/impulsive symptoms than inattention ^21^. In addition, the dysfunction of these affected regions has also been associated with various cognitive deficits in patients with ADHD ^2, 22^.

Studies of adult patients with ADHD, relatively sparse compared to children, showed that adult patients presented impairments in both inattention and hyperactivity domains, but were more affected by inattention than hyperactivity ^23^. Their cognitive ability was also affected heterogeneously ^24, 25^ with working memory impairment being most frequently documented ^3, 26^. GM alterations were overall shown in specific brain regions with the cerebellum and frontal cortex ^19, 27, 28^ being reported more consistently than subcortical regions ^13, 29, 30^. Our previous study has observed significant working memory deficit in adult patients, and it was associated with GM alterations in prefrontal and cerebellar regions ^31^.

Few studies have compared GM anomalies between children, adolescents and adults with ADHD. From studies of pre-selected brain regions, the caudate nucleus, putamen, and amygdala were significantly reduced in patients and the reduction seemed to diminish over time from childhood to adulthood ^12, 13, 32^. The orbital frontal areas were altered in both children and young adults and were related to current and future ADHD symptoms ^33, 34^. Whole brain voxel-based morphometry studies reported additional cortical regions including the anterior cingulate being affected more in adults than children ^13^. Our study focused on GM alterations associated with specific symptom domains and/or working memory deficit, beyond the diagnosis. This work utilized an Independent Component Analyses (ICA) approach that surveys the whole brain without pre-selection of regions of interest and segments the brain into independent components (coherent GM networks). Specifically, GM networks of two age groups, adults and adolescents, were extracted separately, tested for associations with symptoms and working memory in their own group, and then cross evaluated for the other group. Our previous study has analyzed the adult group and reported five GM networks of interest ^31^. Here we extended into the adolescent group and focused on the relationship between the two age groups.

## Participants and methods

### Participants

This study analyzed data from two ADHD projects: the Dutch chapter of the International Multicentre persistent ADHD genetics CollaboraTion (IMpACT) consortium ^25, 35^, and the NeuroIMAGE project ^36^. The IMpACT project recruited adult participants with ADHD and healthy controls, while the NeuroIMAGE project recruited participants from ADHD families (including probands and siblings) and healthy controls in childhood and then followed them up. Data used here were from the 1^st^ MRI scan with some participants in adolescence and some in adulthood. The inclusion and exclusion criteria were described in the original papers ^35, 36^. In brief, the IMpACT ADHD participants met DSM-IV-TR criteria for ADHD in adulthood as well as childhood retrospectively. The NeuroIMAGE participants met DSM-IV-TR criteria for children or adult ADHD, and adult participants also had a formal and research diagnosis in childhood. All participants had IQ≥70, no diagnosis of autism, epilepsy, brain disorders and any genetic or medical disorders related to externalizing behaviors which might be confused with ADHD.

The Dutch chapter of IMpACT study was approved by the regional ethics committee (Centrale Commissie Mensgebonden Onderzoek: CMO Regio Arnhem – Nijmegen; Protocol number III.04.0403). Written informed consent was obtained from all participants. The NeuroIMAGE study was approved by the regional ethics committee (Centrale Commissie Mensgebonden Onderzoek: CMO Regio Arnhem Nijmegen; 2008/163; ABR: NL23894.091.08) and the medical ethical committee of the VU University Medical Center. Written informed consent for every participant was obtained. For children 12–18 years old, both parents and children gave consent, for children younger than 12 parents gave consent for their children.

In order to compare adults with adolescents, the participants of IMpACT and NeuroIMAGE projects were regrouped into adult samples (N=486, age>=18, including participants of IMpACT and NeuroIMAGE) and adolescent samples (N=508, 7<age<18, part of NeuroIMAGE; we named this group as adolescents since 436 participants were older than 12). A summary of participant demographics is shown in Table 1. Concerning the large age range in each group, we also tested subsets of age limited participants, which included 427 adults (18<= age <40, control/case/sibling=139/192/96, female/male=212/215) and 436 adolescents (12<age<18, female/male= 177/259, controls/cases/siblings=137/174/125).

**Table 1.** Demographic and assessment information of participants

|  | Adults (age 18-63) |  |  | Adolescents (age:7-17) |  |  |
| --- | --- | --- | --- | --- | --- | --- |
|  | ADHD | Siblings | Controls | ADHD | Siblings | Controls |
| N /male/<br>medicated | 214/123/105 | 96/49/0 | 176/51/0 | 210/129/121 | 140/59/12 | 158/93/2 |
| Age | 25.35±8.56 | 21.41 ± 2.34 | 28.93 ± 11.79 | 14.61±2.41 | 14.80±2.07 | 14.56± 2.17 |
| IA | 7.23±1.66 | 1.60 ± 1.97 | 0.56 ± 1.24 | 7.33±1.71 | 1.34± 1.96 | 0.77± 1.67 |
| HI | 5.79±2.38 | 1.48±1.65 | 0.70 ± 1.12 | 5.99±2.39 | 0.99± 1.57 | 0.36± 1.10 |
| Digit span<br>forward | 7.94±1.79 | 8.58±1.67 | 8.76±1.73 | 8.86±1.96 | 9.39±1.88 | 9.58±1.94 |
| Digit span<br>backward | 5.09±1.74 | 5.99±1.69 | 6.12±2.02 | 6.45±2.25 | 6.63±2.36 | 7.39±2.05 |
| Scan* | 58/68/88 | 40/56/0 | 47/32/97 | 95/115/0 | 76/64/0 | 95/115/0 |

IA: inattention; HI: hyperactivity/impulsivity; Scan\*: NeuroIMAGE 1/ NeuroIMAGE 2/ IMpACT (Dutch)

### ADHD symptoms and working memory scores

Two ADHD domains, inattention and hyperactivity/impulsivity, were assessed for all participants. NeuroIMAGE used the Schedule for Affective Disorders and Schizophrenia— present and lifetime version, and Conners Teacher Rating Scale −1997 Revised Version: Long Form. IMpACT used the Diagnostic Interview for Adult ADHD. The symptom scores for both domains were counted following the 18 DSM-IV questions, with the score in each domain ranging from 0 to 9. Healthy controls had scores less than 2 in either domain. Unaffected siblings from patient families had scores fewer than 5 or 6 in either domain for adult or adolescent participants, respectively. Both IMpACT and NeuroIMAGE projects conducted WAIS Digit Span test ^37^. We utilized maximum forward and maximum backward scores to gauge working memory capacity (other measures of working memory, such as visual-spatial working memory test for functional MRI, were only available for part of participants. Thus we only focused on digit span test). In the adult group, participants with ADHD had significantly lower scores in both forward and backward tests than controls (p=4.36×10^−4^; p=3.76×10^−5^, respectively). Similarly, adolescent participants with ADHD also presented significantly lower forward and backward scores (p=1.57×10^−4^; p=5.58×10^−6^, respectively).

### Imaging data acquisition and processing

T1-weighted MRI images were acquired with three 1.5T scanners with closely matched settings. Thorough quality control was applied to the imaging data as previously described, including selecting better one from the two sessions. MRI quality indexes (coefficient of joint variation, contrast-to-noise ratio, entropy focus criterion^38^) were generated for further quality check. All good quality images were segmented into six types of tissues using Statistical Parametric Mapping 12 (SPM12, http://www.fil.ion.ucl.ac.uk/spm/software/spm12/), followed by normalization to the Montreal Neurological Institute space, modulated, and smoothed with a 6×6×6 mm^3^ Gaussian kernel. The only difference during the preprocessing of adult and adolescent data was the tissue probability map templates used for segmentation, where SPM12 templates were used for adults and age-specific tissue map templates generated by TOM tool ^39^ were for adolescents. Further analyses on segmented GM images were done separately for adults and adolescents, including selecting individual maps that had correlations with the group mean GM hinger than 0.8, generating a GM mask for each group to include voxels with group mean GM volume larger than 0.2, and regressing out effects of age, sex and site using a linear regression model for each included voxel. The GM mask of adults was slightly different from adolescents, and comparison analyses were applied only to the common voxels (441,258 voxels).

### GM components in adults and adolescents

In the ICA model, **X**= **AS, X** is the GM data matrix, **S** is the component matrix, and **A** is the loading matrix. Each row of S shows weights of individual voxels in one component. Each column of **A** shows expression or loadings of individual subjects for one component. A component reflects coherent GM variations within a brain network with flexibility to go beyond the known boundaries of brain anatomy ^40^. This approach not only reduces multiple comparisons required in whole brain analyses, but also dissects the brain into structurally independent networks. For example, large areas of the frontal lobe may all relate to working memory, but different subregions may relate to working memory in different ways. Whole brain voxel-wise analysis based on association significance would not be able to separate them, while ICA separates brain into independent networks first and then tests each network’s properties.

As reported in our previous study when ICA was applied on adult GM data ^31^, five components out of 22 were significantly related to inattention, working memory deficit, or ADHD diagnosis in adult participants (supplementary text). To test how these structural alterations present in adolescents, we projected these GM components onto the adolescent data. Specifically, using the components derived from adults S, we applied **A**_**c**_=**X**_**c**_**S**^-1^, where **X**_**c**_ was the adolescent GM data, yielding the projected loading matrix **A**_**c**._ **A**_**c**_ showed how the adult brain components were expressed in adolescents, on which we tested the associations with diagnosis, symptoms, and working memory in adolescents. Vice versa, we applied ICA to the adolescent GM data in the same manner. Twenty components were extracted and tested for associations with diagnosis, symptoms, and working memory. For the components with significant associations, we projected them onto the adult data, followed by association tests with diagnosis, symptom and working memory in adults. In addition, due to our specific interest on the caudate nucleus, which has been reported frequently for GM reduction in adolescents with ADHD, we selected specifically components including caudate nucleus from both datasets, and tested their associations with diagnosis, working memory, and symptoms.

### Statistical analyses

A linear mixed model with family structure as a random effect and diagnosis as a fixed effect were applied onto GM loadings of cases and controls to test case vs. control differences (age, sex and site had been regressed out voxel-wise in the preprocess). Similarly, linear mixed models with symptom score or working memory score as a dependent variable, GM loading, age, and sex as fixed effects and family as a random effect were applied to test GM association with symptoms or working memory using all participants (cases, control and siblings) in the group. Given that medication can affect both brain and behavior, we first compared GM loadings between medicated cases and unmedicated cases. If there was a significant difference (p<0.05), we added medication status (used vs. not used) as a fixed effect in the linear models.

Comorbidity with major depression and anxiety were also tested by adding them separately as a covariate into the linear models. We did not control for IQ, because it can remove ADHD related variance ^41^. Note that for both directions of cross evaluation (components discovered in adults→projection to adolescents for evaluation, and components discovered in adolescents→projection to adults for evaluation), we applied false discovery rate (FDR) p<0.05 to control for multiple comparisons (22 components for adults and 20 components for adolescents, respectively) on the discovery results, and uncorrected p<0.05 for evaluation of the projected results.

Given the large age range of participants, particularly in adults, we performed additional analyses to enhance the stability of test results. First, we selected participants with age between 18 and 40 years old, performed a separate ICA and compared the resultant components with those derived from full adult samples reported in ^31^. Second, we replicated association tests with working memory and symptoms for the identified components using homogenous subset of adults(18-40 years old) and adolescents (12-17 years old).

## Results

In the previous study we have reported five GM components in adults significantly associated with working memory, inattention, or diagnosis. The analyses and results of our previous study are summarized in the supplementary text. The five components are plotted in Figure 1 (Independent Component (IC) 1-5). Using the homogenous subgroup 427 participants’ GM data, a separate ICA extracted highly similar components. The correlations between subgroup components and full-sample components were 0.97, 0.85, 0.95, 0.96, and 0.97 for the five components, respectively. The correlations between loading coefficients of the components were 0.98, 0.96, 0.99, 0.98 and 0.98. The validity of the five components has been provided before ^31^. Additionally, we tested possible confounding effects from three MRI quality metrics (coefficient of joint variation, contrast-to-noise ratio, entropy focus criterion), using two regression models: 1) working memory or symptom = GM loading + age +gender + family (random effect) + three MRI quality metrics, and 2) GM loading = case/control + family + three MRI quality metrics. The new test results agreed with previous reported association results (components were significantly associated with working memory, inhibition or case control status with p<0.003). Given the high consistence, we continued our analyses with the full-sample derived components previously reported.

**Figure 1.**
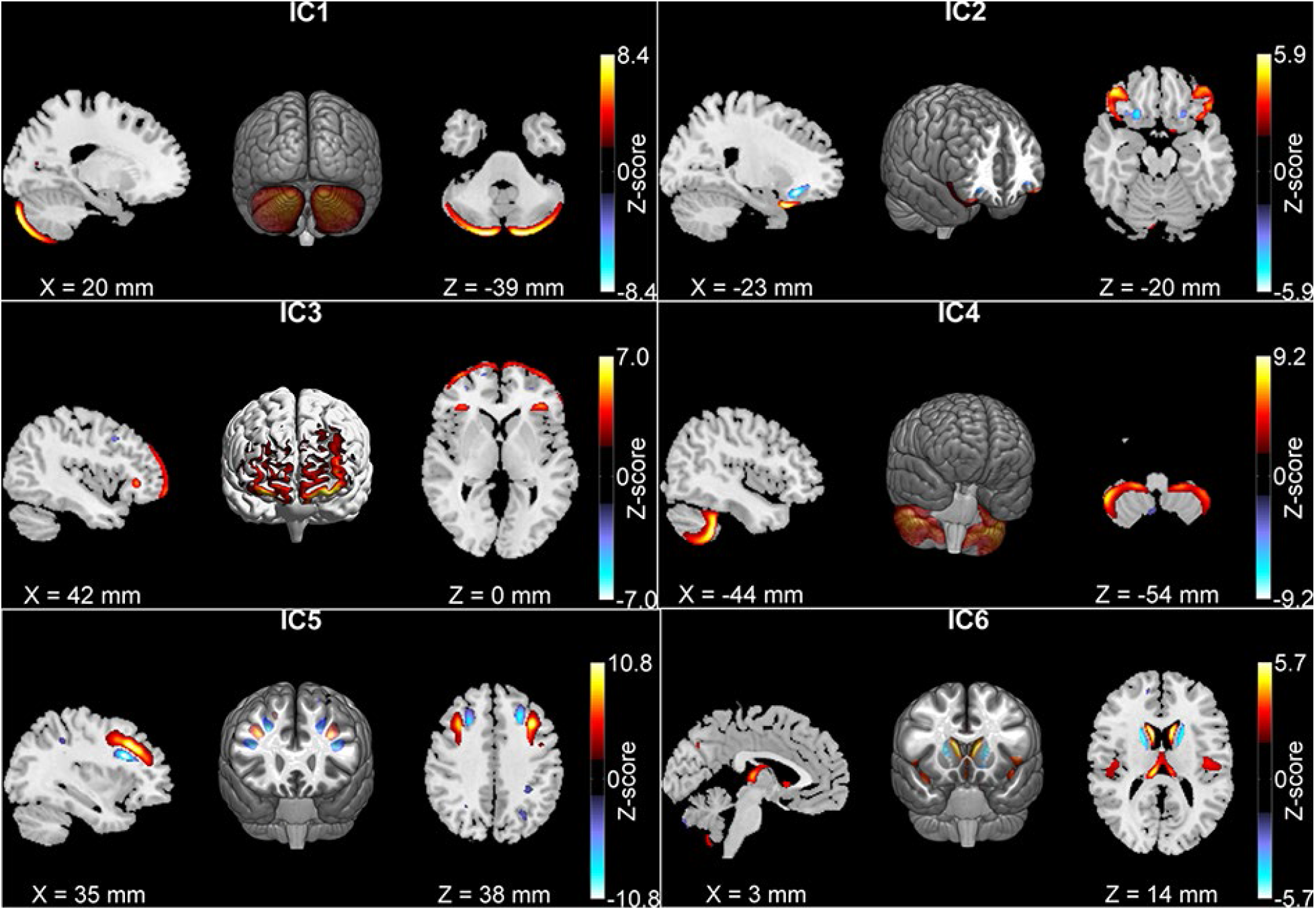
The six gray matter components identified in adults. In adults ICs 1-3 were associated with working memory, IC 4 was associated with inattention, IC 5 showed gray matter reduction in patients, and IC 6 included the caudate nucleus.

After we projected the five components into adolescents, their associations with working memory and symptoms in both adults and adolescents were listed in Table 2A. In adults, Components 1-4 showed no GM differences between medicated and unmedicated cases (p>0.05), while Component 5 showed a significant medication effect. Thus, association results of Component 5 were computed with a linear model controlling for medication. In adolescents, Components 1-4 showed significant GM increases in medicated cases compared to unmedicated cases (p=2.08×10^−2^, 4.43×10^−3^, 1.73×10^−2^, 1.22×10^−2^, respectively). Thus, their association results were from linear models controlling for medication. Component 5 showed no medication effect and no case vs. control difference. No significant comorbidity effect was observed for these components in both adults and adolescents.

**Table 2.** Association results of gray matter components identified in adults and adolescents

| A: GM components identified in the adult group |  |  |  |
| --- | --- | --- | --- |
|  |  | Association in adults | Association in adolescents |
| IC 1 | case vs. control | n.s. | p=2.63×10<sup>-2</sup>; reduction, |
|  | association | **with working memory (forward)**<br>**p=1.4×10<sup>-4</sup>; positive, R<sup>2</sup>=3.08%** | with working memory (forward)<br>n.s. |
| IC 2 | case vs. control | n.s. | p=1.56×10<sup>-4</sup>; reduction, |
|  | association* | **with working memory (forward)**<br>**p=2.68×10<sup>-3</sup>; positive, R<sup>2</sup>=1.90%** | **with working memory (forward)**<br>**p=2.72×10<sup>-2</sup>; positive, R<sup>2</sup>=0.88%** |
| IC 3 | case vs. control | n.s. | p=4.51×10<sup>-3</sup>; reduction, |
|  | association* | **with working memory (backward)**<br>**p=1.66×10<sup>-3</sup>; positive, R<sup>2</sup>= 2.05%** | **with working memory (forward)**<br>**p=1.33×10<sup>-2</sup>; positive, R<sup>2</sup>=1.13%** |
| IC 4 | case vs. control | p=1.04×10<sup>-2</sup>; reduction | p=1.35×10<sup>-2</sup>; reduction |
|  | association* | **with inattentive symptom**<br>**p=2.26×10<sup>-3</sup>, negative, R<sup>2</sup>=1.88%** | **with inattentive symptom**<br>**p=3.04×10<sup>-2</sup>, negative, R<sup>2</sup>=0.61%** |
| IC 5 | case vs. control | **p=1.51×10<sup>-4</sup>; reduction** | n.s. |
|  | association | n.s. | n.s. |
| IC 6 | case vs. control | n.s. | p=3.34×10<sup>-2</sup>; reduction |
|  | association | n.s. | with working memory (forward),<br>p=7.86×10<sup>-3</sup>, positive, R<sup>2</sup>=1.34% |
| B: GM components identified in the adolescent group |  |  |  |
|  |  | Association in adults | Association in adolescents |
| IC A | case vs. control | n.s. | **p=5.37×10<sup>-4</sup>; reduction,** |
|  | association* | **with inattentive symptom**<br>**p=2.15×10<sup>-2</sup>; negative, R<sup>2</sup>=1.09%,** | **with inattentive symptom**<br>**p=2.01×10<sup>-3</sup>, negative, R<sup>2</sup>=1.19%** |
| IC B | case vs. control | n.s. | **p=1.78×10<sup>-3</sup>; reduction,** |
|  | association* | **with working memory (forward)**<br>**p=4.58×10<sup>-3</sup>; positive, R<sup>2</sup>=1.71%** | **with working memory (forward)**<br>**p=4.02×10<sup>-3</sup>; positive, R<sup>2</sup>=1.50%** |

n.s. = not significant; Results in bold font are significant, where the significance in discovery data was set to pass FDR p<0.05, and the significance in verification data after projection was set to pass uncorrected p<0.05. p values shown in the table are uncorrected. R<sup>2</sup> indicates variance of working memory or inattention score explained by the IC. \* indicates consistent associations across adolescents and adults.

Across adults and adolescents, three GM components (ICs 2-4 in Table 2A) showed consistent associations with either working memory or inattention. Component 2, the inferior frontal gyrus, and Component 3, the superior and middle frontal gyri, were positively associated with working memory in both adults and adolescents. More GM volume was associated with higher (better) working memory scores in all participants. Component 4, the cerebellar tonsil and culmen, was negatively associated with inattentive symptom in both adults and adolescents, where lower GM volume was associated with higher (worse) inattentive score. In addition to these significant associations, it is noteworthy that adolescent patients showed nominal GM reduction in four of the five components (p<0.05), while adult patients showed GM reduction in two components (one passed FDR and one had p<0.05).

Twenty components were extracted from the adolescent data of which twelve were highly similar to those in adults (r≤0.5, see Supplementary Table 1). Two components (Figure 2) showed significant GM reduction in patients and were also significantly associated with inattention or working memory in adolescents. For these two components, medicated cases had increased GM volumes compared to unmedicated cases (p=4.64×10^−3^ and p=3.95×10^−2^, respectively). After projection to adults, no medication effects were observed. Thus, medication effect was controlled only for adolescents. As shown in Figure 2, Component A comprised left hemisphere, cerebellar tonsil and culmen, lingual gyrus, and cuneus. Component B comprised bilateral insula, inferior frontal, superior temporal gyri, and caudate nucleus. Table 2B lists out associations of the two components with working memory, inattention, and diagnosis in both age groups. GM volume of Component A was negatively associated with inattentive symptom in adolescents and adults and showed significant reduction in adolescents with ADHD. GM volume of Component B was positively associated with working memory in both groups and showed significant reduction in adolescents with ADHD. No significant comorbidity effect was observed for the components in both groups.

**Figure 2.**
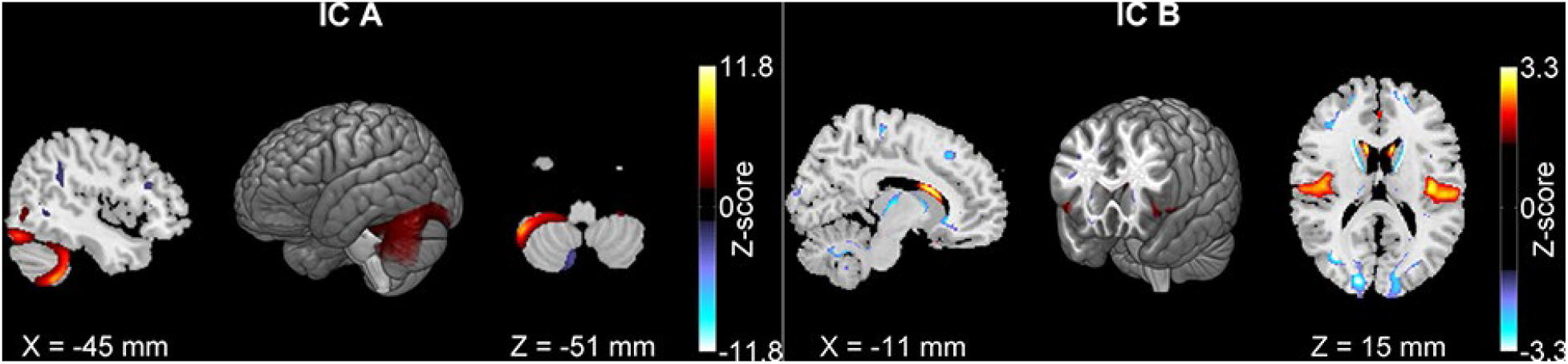
The two gray matter components identified in adolescents. IC A was associated with inattention and IC B was associated with working memory. Both components had gray matter reduction in ADHD patients.

Focusing on the components specifically including the caudate nucleus, we identified one adult component, (Component 6 in Figure 1, comprising caudate nucleus, superior temporal and insula), and one adolescent component (Component B in Figure 2). These two components were highly correlated (r=0.58, p<1e-16). But the adolescent component had more areas in insula and inferior frontal gyrus and less in the caudate nucleus relative to the adult one. As listed in Table 2A, Row IC 6 and Table 2B, Row IC B, the adult component showed none associations in adults but nominal associations with working memory and diagnosis in adolescents (p<0.05), while the adolescent Component B showed consistent significant associations with working memory in both groups and significant GM reduction in adolescents.

Due to the large age range of each group, we replicated association tests for these eight components using relatively homogenous group settings (adolescents:12-17 years old with 436 participants; young adults: 18-40 years old with 427 participants). The results (supplementary table s2) were in general consisted with those derived using all participants. All associations reported in Tables 2-3 except two presented consistent significant results. The two in discrepancy were where subsamples could not report associations with p<0.05.

We also compared GM volumes of the unaffected siblings with healthy controls and patients with ADHD. Figure 3 plots GM loadings of each diagnostic group, each age group and each component. In adolescents, seven components (except IC5) had some levels of GM reduction in patients with ADHD (significant or nominal, see Table 2). Siblings had more GM volume than patients but less than controls in five components. However, most of the differences were not statistically meaningful (p>0.05). In adults, siblings presented no clear patterns. Similar results were observed when we examined only unmedicated patients (see results of unmedicated cases, siblings, and controls in supplementary Figure s1).

**Figure 3.**
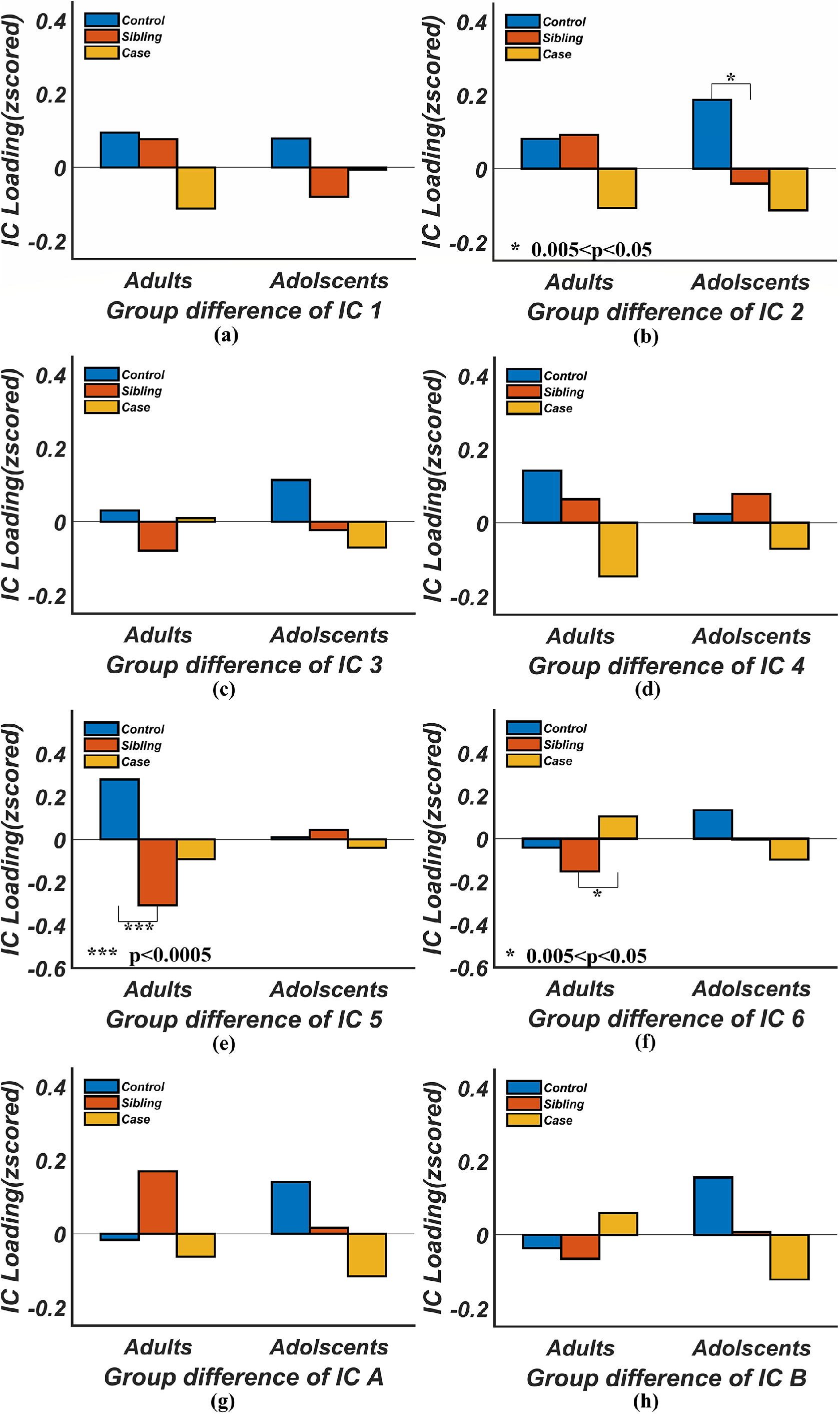
Comparison of gray matter volumes in unaffected siblings against those in controls and patients. P values for the comparisons between cases and controls were listed in Table 2, and not plotted here for the clarity of the figure. Only the significant results for comparisons involving siblings (p<0.05) are plotted.

## Discussions and conclusions

Aiming at the GM alterations of ADHD patients in adolescence and adulthood, we compared GM patterns of the two age groups extracted by data-driven whole-brain ICA approaches. GM networks derived by ICA are sensitive to data characteristics (i.e. intrinsic to the specific cohort). In the twenty components extracted from adolescents, twelve were highly similar to those in adults, supporting continuation of brain structural segmentations. The differences between similar components and the totally different components likely indicate developmental effects on similar regions of brain and the unique constellation to each group. To ensure fair comparisons between the two age groups, we conducted the network discovery-and-verification two-step approach in both directions: from adults to adolescents and from adolescents to adults. Additionally, adolescent patients with ADHD will inevitably include both those likely to persist and those likely to remit. This mixture will most likely contribute to the different GM patterns between the adolescent group and the adult group which included only persistent ADHD patients. Thus, our discussion focuses on consistent findings between the two age groups.

For the four components associated with working memory (three adult components and one adolescent component passing multiple comparison correction), three components showed consistent associations across childhood to adulthood, including the superior, middle, and inferior frontal regions (adult IC 2-3), and the insula, inferior frontal, superior temporal gyri and caudate (adolescent IC B). The prefrontal regions are well-documented for their role in working memory as a central executive point ^42^, involving integrating sensory information, allocating neural resources, maintaining and shifting attention, and updating and manipulating information ^43-45^. Better working memory performance has been found to correlate with greater GM volume in widespread brain areas including the superior, middle, and inferior frontal gyri ^46, 47^. Interestingly, a recent large sample study (1336 young adults) on the relationship between regional GM and cognition reported that the strongest association between GM and working memory was in the insula, which was the only region significantly associated with working memory after controlling for total brain volume ^47^. Our results further suggest these regions are consistently involved in working memory from childhood to adulthood.

Working memory deficits in ADHD have been repeatedly and consistently reported for both adolescents and adults ^1, 26, 48, 49^. The longitudinal study led by Biederman et al. suggested cognitive impairments in ADHD originating in childhood persisted into adulthood ^50^. Our results suggest that not only working memory deficits persisted from childhood and adulthood, also did the neural correlates of working memory deficits, highlighting the frontal regions and insula.

Two components (adult component 4 and adolescent component A) were significantly associated with inattention across both age groups, with more GM volume related to lower inattentive symptom scores. Component 4 mainly consisted of bilateral cerebellar tonsil and culmen, and Component A consisted of left hemisphere, cerebellar tonsil and culmen, lingual gyrus, and cuneus. These two components were spatially similar with a correlation of 0.49, overlapping mostly in left hemisphere cerebellar regions. The differences likely reflect different developmental trajectories of left and right cerebellar hemispheres in adolescence. Both adult and adolescent components support the involvement of cerebellum in ADHD, consistent with repeatedly reported cerebellum volume reduction as the most stable brain alterations observed in ADHD patients ^17, 19, 20, 51-53^.

The role of the cerebellum in cognitive function has been recognized ^54, 55^, supported by cognitive impairment in patients with cerebellum lesions ^56^, prevalent activation during cognitive functions ^57^, anatomic connections between cerebellum and cortical regions ^58, 59^, and functional co-organization with cortical regions ^60, 61^. In ADHD patients, the functional impact of cerebellum GM alteration is less studied, with the potential to affect neurological soft signs ^62, 63^, attention process as part of executive function ^55^, or both as these two could be linked ^64, 65^. Our findings add to the literature and emphasizes a specific role of the cerebellum in the attention process, and its persistence from childhood to adulthood.

A detailed look at the components including the caudate nucleus appeared to show that some caudate areas were integrated together with the insula, inferior frontal, and superior temporal regions for both adults and adolescents. An additional analysis using GM data without voxel-wise regression of age effect extracted a more complete/dominate caudate component (supplementary Figure s2). The spatial difference of the components with and without voxel-wise age regression indicates that age had a significant role and carried a large proportion of variance in the caudate nucleus. In fact, one previous study using NeuroIMAGE data has reported developmentally sensitive caudate alterations in relation to ADHD ^12^: caudate volume reduction in younger ADHD patients (age 8-15), not in middle age patients (age 15-22), and reversed in older patients (age 22-30). Our age grouping and processing approach resulted in the components that included partial caudate and interrelated insula ^66^ and cortical regions ^67^, not a typical whole caudate region. Nevertheless, the comparison of the two components that include caudate nucleus advocates the role of insula and inferior frontal regions in working memory processes, as more consistent and more significant associations with working memory were observed for the component with more insula and inferior frontal region.

In contrast to the consistent results discussed above, we found that GM reductions observed in adolescent ADHD patients were largely not replicated in the adult group. The two components, IC A and IC B showed significant GM reductions in adolescent patients (passing multiple comparison correction), but not in adults. The other five components ICs 1-4 and 6 showed nominal (p<0.05) GM reductions in adolescent patients, and only one component (IC 4) showed comparable levels of reduction in adults. Moreover, we also observed more prevalent medication effects on GM in adolescents than adults. Among the seven components with nominal or significant GM reduction in adolescents, six showed that medication mitigated the reduction in patients. Altogether these findings agree with previous studies reporting GM reduction in ADHD patients diminished with age, particularly in subcortical regions ^13^. Our results expand the affected regions into the frontal and cerebellum regions. One note is that medication could, at least, partially contribute to the observed diminished GM reduction with age.

Comparison of GM volumes of siblings against cases and controls in Figure 3 produced mostly no significant differences. We think this might be due to two reasons. One is that our sample size is too small to reach significant conclusion. The other, which is more likely, is the heterogeneity of the sibling group, whose ADHD symptom scores ranged from 0 to 5/6 (adult/children, respectively). Studies have shown participants with symptom score higher than 2 could be considered as subthreshold ADHD ^68, 69^, and they were at greater risk for negative outcomes in cognitive domains ^70^. Little is known about subthreshold ADHD for which more investigations are needed.

Findings of this study should be interpreted with consideration of the following limitations. First, the age ranges of both groups are large. To mitigate the heterogeneity, we have tested relatively homogenous age groups (adolescents of 12-17 years old and adults of 18-40 years old). Similar findings as discussed above were observed. Second, our analytical approaches focus on the comparison of two age groups with age regression specific to each group. This approach leads to common patterns across the age range within each group, suitable for group comparisons. However, the dynamic developmental trajectories, like the one in the caudate nucleus, are missed. Third, ICA segments brain regions with coherent patterns together, including those with opposite patterns as shown in blue in Figures 1 and 2. The blue regions were smaller and contributed with less weights than the positive regions in red. They showed opposite effects to the positive regions as discussed above. The interpretation of these regions is limited and further specific studies to verify these regions effect are necessary. Finally, even though we have tried to control for confounding factors like medication and comorbidity (depression and anxiety) in the analyses, other factors potentially affecting ADHD, such as obsessive-compulsive disorder and parenting strategies, were not exclusively investigated. The mixture of adolescent patients who likely remit or persist in the future also limits our power to discover the GM patterns associated with disease persistence. Future investigations leveraging longitudinal information of patient disease progression are necessary to verify and refine currently findings.

Overall, through direct comparisons of GM components in both adolescents and adults, our findings suggest that brain regions associated with deficits in working memory and attention in ADHD patients persist from childhood into adulthood. These regions include the inferior/superior/middle frontal gyri and the insula for working memory, and the cerebellar tonsil and culmen for attention. They could be the target for further functional investigation of persistent ADHD. In contrast, typical GM reduction observed in younger patients (adolescents) largely diminishes in adulthood, and this phenomenon is beyond subcortical regions to cortical regions.

## Data Availability

No new data were collected in this study.

## Acknowledgement

We would like to thank the supporting agency of the study, the National Institutes of Health, through the grant 1R01MH106655. This study makes use of data from the Dutch NeuroIMAGE project and the Dutch site of IMpACT (International Multi-center persistent ADHD Collaboration) project. The NeuroIMAGE project was supported by NWO Large Investment grant 1750102007010 (Dr Buitelaar), ZonMW Addiction: Risk Behaviour and Dependency Grant 60-60600-97-193 (Dr Buitelaar), NWO Brain & Cognition: an Integrative Approach grant 433-09-242 (Dr Buitelaar), NWO National Initiative Brain & Cognition 056-13-015 (Dr Buitelaar), the EU FP7 grants TACTICS (278948), IMAGEMEND (602450), MATRICS (603016) and AGGRESSOTYPE (602805), EU IMI grant EU-AIMS (115300) and EU H2020 grant Eat2beNICE (728018) and grants from Radboudumc, University Medical Center Groningen, Accare, and VU University Amsterdam. The Dutch IMpACT study acknowledges the following sources of support: The Netherlands Organization for Scientific Research (NWO), i.e. the NWO Brain & Cognition Excellence Program (grant 433-09-229) and the Vici Innovation Program (grant 016-130-669 to BF). Additional support was received from the European Community’s Seventh Framework Programme (FP7/2007 – 2013) under grant agreements n° 602805 (Aggressotype), n° 602450 (IMAGEMEND), and n° 278948 (TACTICS) as well as from the European Community’s Horizon 2020 Programme (H2020/2014 – 2020) under grant agreements n° 643051 (MiND), Eat2beNICE (728018) and n° 667302 (CoCA). The work was also supported by grants for the ENIGMA Consortium (grant number U54 EB020403) from the BD2K Initiative of a cross-NIH partnership, and by the ECNP Network ADHD across the Lifespan. The authors also thank all participants to these two projects.

